# Variations in COVID-19 impacts by social vulnerability in Philadelphia, June 2020-December 2022

**DOI:** 10.1101/2023.01.13.23284488

**Authors:** Katherine M. Strelau, Rachel Feuerstein-Simon, Nawar Naseer, Megan Lang, Kevin Rix, Eleanor J Murray, Jeffrey S. Morris, Douglas J. Wiebe, Hillary C.M. Nelson, Ronald G. Collman, Kyle G. Rodino, Frederic D. Bushman, Brendan J. Kelly, Carolyn C. Cannuscio

## Abstract

**Introduction:** The study objective was to elucidate the relationship between social vulnerability and COVID-19 impacts in Philadelphia over a 2.5-year period, between June 2020 and December 2022.

**Methods:** Using publicly available COVID-19 case, test, hospitalization, and mortality data for Philadelphia (June 7, 2020-December 31, 2022) and area-level social vulnerability data, we compared the incidence, test positivity, hospitalization, and mortality rates in high and low vulnerability neighborhoods of Philadelphia, characterized as scoring above or below the national median score on the social vulnerability index. We used linear mixed effects models to test the association between social vulnerability and COVID-19 incidence, test positivity, hospitalization, and mortality rates, adjusting for time and age distribution.

**Results:** 90.4% of Philadelphians (*n* = 1,430,153) live in neighborhoods classified as socially vulnerable, based on scoring above the national median score on the social vulnerability index. COVID-19 incidence, hospitalization, and mortality rates were significantly elevated in the more vulnerable communities, with *p* < 0.05, *p* < 0.005, and *p* < 0.001, respectively. The relative risks of COVID-19-related incidence, hospitalization, and death, comparing the more vulnerable neighborhoods to the less vulnerable neighborhoods, were 1.11 (95%CI: 1.10-1.12), 2.07 (95%CI: 1.93-2.20), and 2.06 (95%CI: 1.78-2.38), respectively. Thus, between June 7, 2020 and December 31, 2022, 32,573 COVID-19 cases, 9,409 hospitalizations, and 1,967 deaths would have been avoided in Philadelphia’s more vulnerable communities had they experienced the same rates of incidence, hospitalization, and death as the less vulnerable Philadelphia communities.

**Conclusions:** These results highlight the disparate morbidity and mortality experienced by people living in more vulnerable neighborhoods in a large US city. Importantly, our findings illustrate the importance of designing public health policies and interventions with an equity-driven approach, with greater resources and more intensive prevention strategies applied in socially vulnerable communities.

## Introduction

Officially, over 100 million cases of COVID-19 have been documented across the United States as of December 19, 2022^1^. However, estimates suggest that the true cumulative incidence is far higher, and that almost 2 in 3 Americans have been infected at least once with SARS-CoV-2^2^. The official mortality toll in the US, which was roughly 1.1 million deaths by the end of 2022, may also be an underestimate, given that there have been 1,261,192 excess deaths nationwide between February 2020 - January 2023^3^. Furthermore, the impact of the COVID-19 pandemic has been unequal across populations. COVID-19 case, hospitalization, and death rates among Black and Hispanic populations have outpaced those among White populations^4,5^. There is evidence that COVID-19-related morbidity and mortality may disproportionately impact vulnerable communities^6,7^. Using data from Philadelphia, which is the poorest large city in the United States^8^, we report on the disparate burden of COVID-19 across neighborhoods characterized by higher and lower levels of social and economic vulnerability.

## Methods

### Assessment of social vulnerability and population characteristics

The CDC defines social vulnerability as “the potential negative effects on communities caused by external stresses on human health”^9^. Operationally, we classified Philadelphia’s neighborhoods as areas of higher or lower social vulnerability, using the CDC’s validated Social Vulnerability Index (SVI). This index measures vulnerability in four domains: socioeconomic status, household composition/disability, minority status/language, and housing/transportation^9^. The SVI was developed as a tool for identifying areas in need of enhanced assistance following public health emergencies. We employed the SVI as a composite measure of social and economic disadvantage to assess whether COVID-19 burdens were unequally borne by communities across the socioeconomic spectrum.

The CDC used American Community Survey (ACS) data from 2014-2018 to assign an SVI score (range: 0-1) to each census tract in the United States. Higher scores indicated higher levels of social vulnerability. Because Philadelphia’s COVID-19 test positivity, incidence, hospitalization, and mortality data (our endpoints) were reported by ZIP code, we converted SVI data (our exposure of interest) from the census tract level to the ZIP code level using ArcGIS (v10.7.1) (ESRI, Chesterbrook, PA). We merged the ZIP code and census polygons and calculated the average SVI score across all census tracts in that ZIP code. We then classified Philadelphia’s 46 residential ZIP codes into two groups, based on the national median SVI score (0.5): neighborhoods above the national median SVI score (i.e., Philadelphia neighborhoods more vulnerable than 50% of census tracts in the United States) and neighborhoods below the national median SVI score (i.e., neighborhoods less vulnerable than 50% of census tracts in the United States). Throughout the paper, we refer to Philadelphia’s “more vulnerable” and “less vulnerable” communities based on this nationally relevant cut-point.

Age and race distributions were based on the 2020 5-year ACS^10^. The results were plotted in Prism (v9.4.1) (GraphPad Software, San Diego CA).

### Assessment of COVID-19 disease burden

We assessed the local burden of COVID-19 using publicly available data to identify weekly COVID-19 incidence, hospitalization, mortality, and test positivity within each ZIP code in Philadelphia County, whose boundaries are contiguous with those of the city of Philadelphia. We used daily case, test, hospitalization, and death counts reported on Open Data Philly to create individual datasets for each metric per ZIP code on a weekly basis. Based on data availability, weekly case, test, and death counts are reported between June 7, 2020 and December 31, 2022, and hospitalization counts are reported between August 16, 2020 and December 31, 2022, subsequently referred to as the study period. We then combined the individual datasets for weekly case, test, hospitalization, and death counts with population based on the 2020 5-year ACS^10^ and SVI category (above or below the national median SVI by ZIP code).

### Statistical Analyses

We calculated the crude (unadjusted) weekly and cumulative incidence, hospitalization, and mortality rates per 100,000 residents as well as the test positivity rate for each ZIP code and then plotted the results using Prism. We also calculated the relative risk of confirmed COVID-19 incidence, hospitalization, or mortality rates by social vulnerability, and the corresponding 95% confidence intervals (**Fig. 3, middle**).

To determine the number of cases, hospitalizations, and deaths that would have been avoided if the more vulnerable communities had experienced the same rates as the less vulnerable communities, we calculated the cumulative incidence, hospitalization, and mortality rates for the less vulnerable communities throughout the duration of the study period. These rates were then multiplied by the population denominator in the more vulnerable communities to find the predicted number of cases, hospitalizations, and deaths that would have occurred if the more vulnerable communities had experienced the same rates as the less vulnerable communities. These numbers were then subtracted from the total number of actual cases, hospitalizations, and deaths that had been recorded in the more vulnerable neighborhoods.

We used linear mixed models in R (using the function lmer() from the lme4 package v1.1-31 created to examine the association between SVI and COVID-19 morbidity and mortality^11^. Multivariate models adjusted for time and population age distribution.

## Results

### A majority of Philadelphians live in ZIP Codes with high social vulnerability

Figure 1 shows characteristics of Philadelphia residents. Of the city’s 46 residential ZIP codes, 38 had an SVI measure above the national median (indicating higher area-level vulnerability), and eight had an SVI measure below the national median (indicating lower area-level vulnerability) (**Fig 1A**). This means that 1,430,153 (90.4%) Philadelphians live in ZIP codes classified as “more vulnerable,” based on the national median SVI, while only 151,595 (9.6%) Philadelphians live in ZIP codes that would be classified as “less vulnerable” by that nationally relevant cut-point (**Fig 1B**).

**Figure 1.**
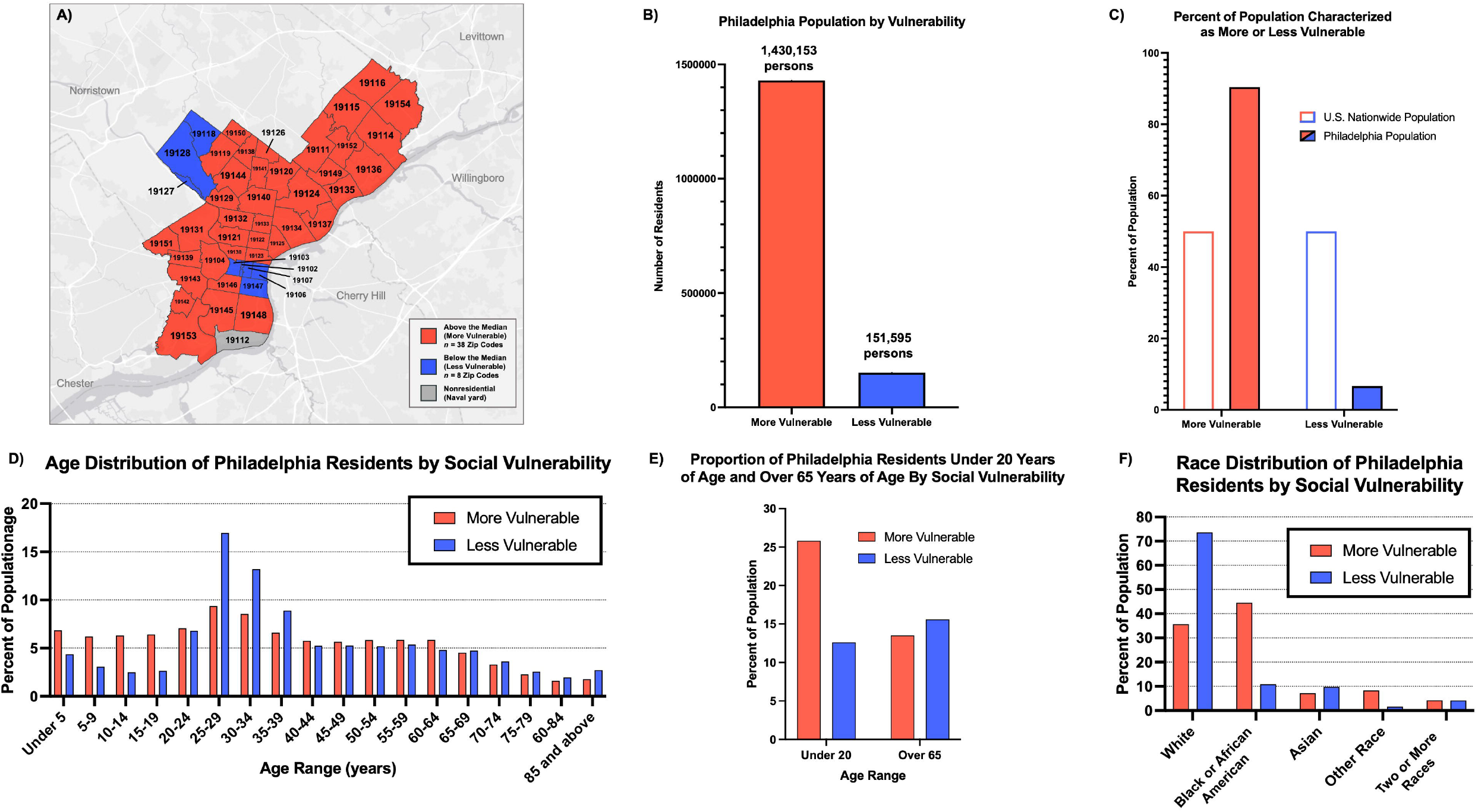
Characteristics of Philadelphians living in more and less vulnerable ZIP codes (n = 46). Less vulnerable zip codes (blue, n = 8) have an SVI below the national median. More vulnerable ZIP codes (red, n = 38) have an SVI above the national median. **A)** Geographical map of Philadelphia ZIP codes indicating predominance of vulnerable communities. **B)** Philadelphia residential population plotted based on vulnerability. **C)** Philadelphia and the US Nationwide populations plotted based on vulnerability. 90.4% of Philadelphians live in ZIP codes characterized as being more vulnerable, and 9.6% of Philadelphians live in ZIP codes characterized as being less vulnerable. **D)** Age distribution of Philadelphia residents by social vulnerability. **E)** Proportion of Philadelphia residents under 20 years of age and over 65 years of age by social vulnerability. **F)** Race distribution of Philadelphia residents by social vulnerability. “Other Race” includes American Indiana or Alaskan Native, Native Hawaiian or Pacific Islander, and Other categories from the US census.

### More vulnerable communities experienced higher pandemic burden than less vulnerable communities

During the study period, COVID-19 incidence appeared to differ most markedly between more and less vulnerable communities at three key points; between November 2020 and January 2021; between March 2021 and May 2021; and between March 2022 and July 2022 (**Fig. 2A**). These time periods corresponded with the fall 2020 holiday season, the Alpha wave, and the Omicron BA.2 wave, respectively (with the variant-specific waves determined using the online dashboard published by Everett et al., 2021^12^). During the fall 2020 holiday season and the Alpha wave, the incidence rate was higher in the more vulnerable communities compared to the less vulnerable communities. During the Omicron BA.2 wave, the incidence rate was higher in the less vulnerable communities compared to the more vulnerable communities, notably for the first time throughout the pandemic. A linear mixed model controlling for time indicated a significantly positive association between higher vulnerability and incidence rate (*p* = 0.0338), with an average of 28.40 more cases per 100,000 per week occurring in the more vulnerable neighborhoods (**Table 1**).

**Table 1.** Summary of the linear mixed models examining the association between each COVID-19 epidemiological rates and vulnerability, controlling for time and age. ß = Estimate (beta), SE = Standard Error, Sig = Significance. ns = no significance, * = p<0.05, ** = p<0.01, ***p<0.001.

| Outcome Variable | Predictor Variable (SVI) | $\beta$ | SE | t value | p value | Sig | Interpretation |
| --- | --- | --- | --- | --- | --- | --- | --- |
| **Incidence Rate** | More Vulnerable | 28.40 | 12.90 | 2.21 | 0.0338 | * | On average, 28.40 more cases per 100,000 per week occurred in the more vulnerable neighborhoods compared to the less vulnerable neighborhoods |
| **Positivity Rate** | More Vulnerable | -0.35 | 0.94 | -0.37 | 0.71 | ns | NA |
| **Hospitalization Rate** | More Vulnerable | 4.84 | 1.48 | 3.26 | 0.0024 | ** | On average, 4.84 more hospitalizations per 100,000 per week occurred in the more vulnerable neighborhoods compared to the less vulnerable neighborhoods |
| **Mortality Rate** | More Vulnerable | 1.47 | 0.39 | 3.77 | 0.000594 | *** | On average, 1.47 more deaths per 100,000 per week occurred in the more vulnerable neighborhoods compared to the less vulnerable neighborhoods |

**Figure 2.**
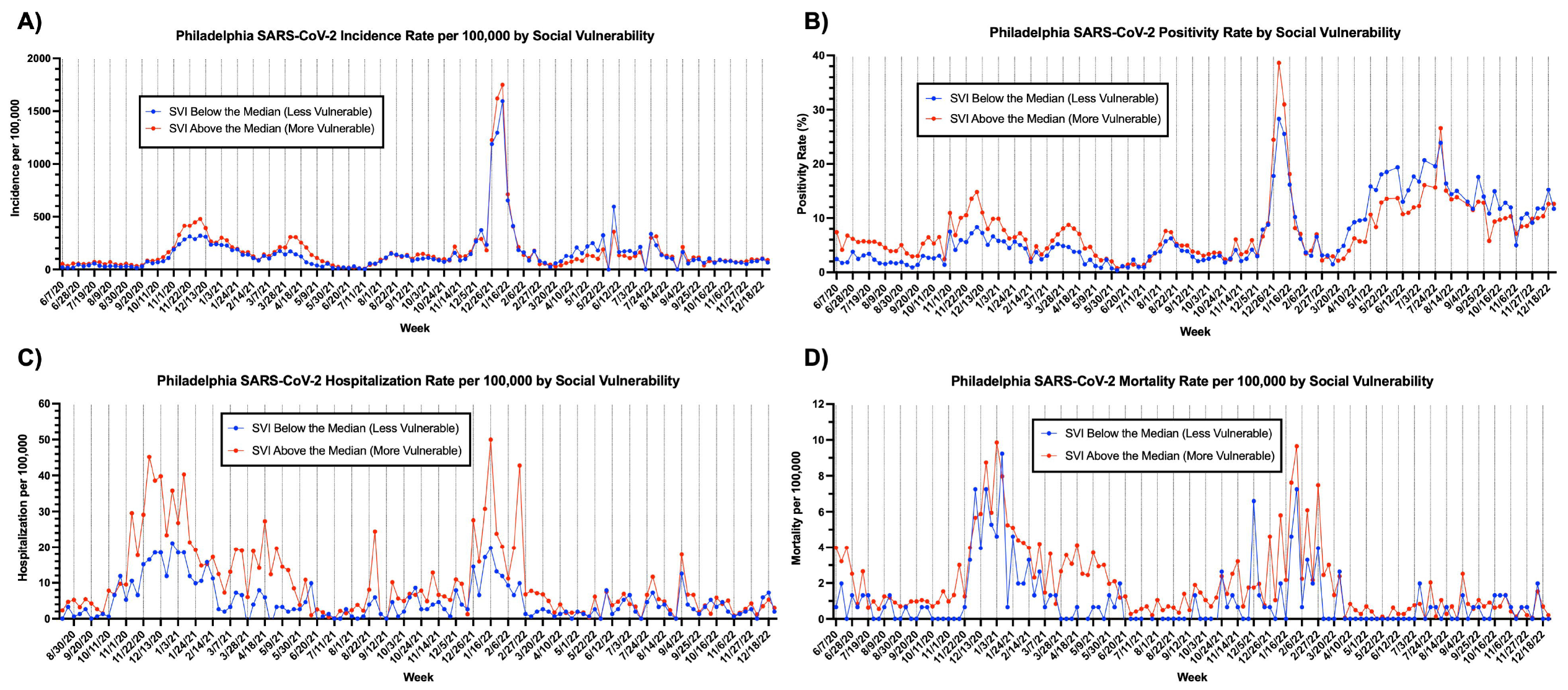
Descriptive metrics of SARS-CoV-2 transmission in Philadelphia by week (June 7, 2020 – Dec 31, 2022) indicate that more vulnerable communities experienced higher rates compared to less vulnerable communities. Less vulnerable communities (blue) had an SVI below the national median. More vulnerable communities (red) had an SVI above the national median. Every third week date is listed on the x-axis for legibility. **A)** Incidence rate of COVID-19 per 100,000 between 06/07/2020 – 12/31/2022 by social vulnerability. **B)** Positivity rate of COVID-19 as a percent between 06/07/2020 – 12/31/2022 by social vulnerability. **C)** Hospitalization rate of COVID-19 per 100,000 between 08/16/2020 – 12/31/2022 by social vulnerability. **D)** Mortality rate of COVID-19 per 100,00 between 06/07/2020 – 12/31/2022 by social vulnerability.

Throughout the study period, the test positivity, hospitalization, and mortality rates were generally higher in the more vulnerable communities compared to the less vulnerable communities **(Fig. 2B-D**). The exception to this observation was the positivity rate between March 20, 2022 and July 24, 2022, which roughly corresponds with the Omicron BA.2 wave. During this time period, the test positivity rate was higher in the less vulnerable communities compared to the more vulnerable communities (**Fig 2B**).

To examine the associations between vulnerability and test positivity, hospitalization, and mortality rate, we built multivariate models that controlled for time, adjusted for age, and treated SVI as a dichotomous predictor variable (high vs. low vulnerability). Hospitalization and mortality rates were significantly positively associated with social vulnerability (*p* = 0.0024 and *p* = 0.0006, respectively), with an average of 4.84 more hospitalization per 100,00 per week and 1.47 more deaths per 100,000 per week occurring in the more vulnerable neighborhoods (**Table 1**).

### Cumulative toll of COVID-19 was worse in more vulnerable communities

To examine the impact of the COVID-19 pandemic among Philadelphians, we calculated the cumulative incidence, hospitalization, and mortality in communities classified as more and less vulnerable (i.e., above and below the national median SVI score) between June 7, 2020 – December 31, 2022, August 16, 2020 – December 31, 2022, and June 7, 2020 – December 31, 2022, respectively. For each measure of disease burden, the impact over that time period was worse in the more vulnerable communities compared to the less vulnerable communities (**Fig. 3A-C, left**).

**Figure 3.**
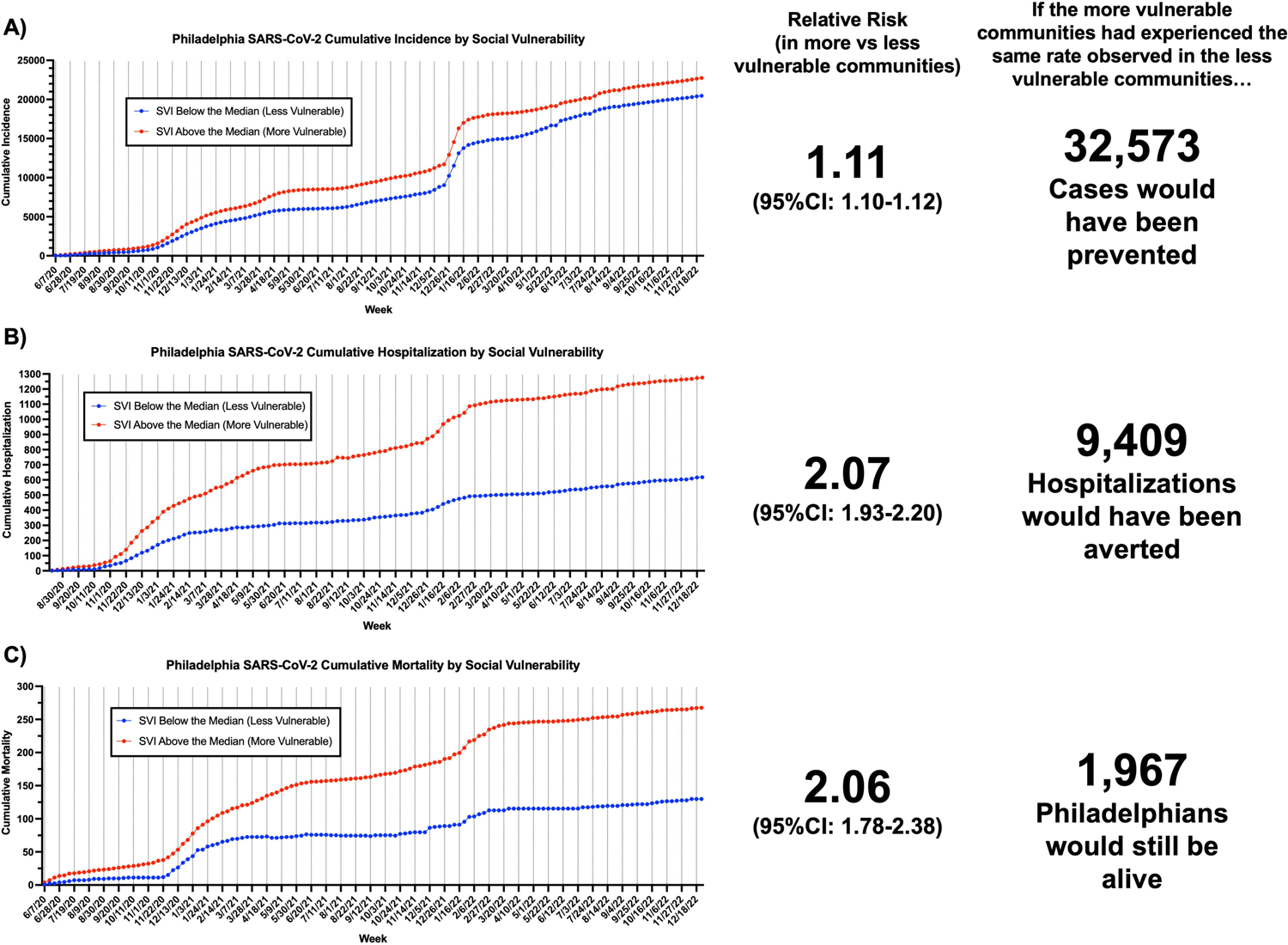
The cumulative impact of the COVID-19 pandemic (June 7, 2020 – Dec 31, 2022) was greater in more vulnerable communities compared to less vulnerable communities. **Left:** Plots for each cumulative metric. Less vulnerable communities (blue) had an SVI below the national median. More vulnerable communities (red) had an SVI above the national median. Every other week date is listed on the x-axis for legibility. **Middle:** Relative risk of each metric (incidence, hospitalization, mortality) in more vs less vulnerable communities. **Right:** The estimated number of cases, hospitalizations, deaths that would have been prevented in the more vulnerable communities if these communities had the same rates as the less vulnerable communities. **A)** Cumulative incidence of SARS-CoV-2 per 100,000 between 06/07/2020 – 12/31/2022. **B)** Cumulative hospitalization rate of SARS-CoV-2 per 100,000 between 08/16/2020 12/31/2022. **C)** Cumulative mortality of SARS-CoV-2 per 100,00 between 06/07/2020 – 12/31/2022.

To better understand the differences in these impacts by area-level social vulnerability, we calculated the relative risk of each cumulative measure in the more vulnerable communities compared to the less vulnerable communities. These relative risks were 1.11 (95%CI: 1.10-1.12), 2.07 (95%CI: 1.93-2.20), and 2.06 (95%CI: 1.78-2.38) for incidence of confirmed cases, hospitalization, and mortality, respectively (**Fig 3A-C, middle**).

Furthermore, we estimated the number of cases, hospitalizations, and deaths that occurred in the more vulnerable communities that would have been prevented if these communities had experienced the same rates as the less vulnerable communities. During the period from June 7, 2020 – December 31, 2022, if the more vulnerable communities had experienced the same incidence of confirmed COVID-19 as the less vulnerable communities, 32,572 fewer persons living in more vulnerable communities would have had confirmed cases of COVID-19 (**Fig. 3A, right**). If the more vulnerable communities had experienced the same hospitalization rate as the less vulnerable communities, there would have been 9,409 fewer persons hospitalized in the more vulnerable communities than were hospitalized between August 16, 2020 – December 31, 2022 (**Fig. 3B, right**). Finally, if the more vulnerable communities had experienced the same mortality rate as the less vulnerable communities from June 7, 2020 – December 31, 2022, 1,967 people would still be alive in the more vulnerable communities today (**Fig. 3C, right**).

## Discussion

These results from Philadelphia advance discussion regarding COVID-19 health inequities by offering insights from the poorest of our nation’s large cities. Results from this study should be compared to findings from other cities across the US, to determine whether and to what degree pandemic inequities persist across different populations and places, and in the context of different pandemic policy responses. In Philadelphia, unequal pandemic burdens were striking, and the cumulative toll of COVID-19, through the end of 2022, was elevated—in terms of confirmed cases, hospitalization, and mortality—in neighborhoods characterized by high levels of social vulnerability (**Figs 2-3, Table 1**). In Philadelphia, 90.4% of residents live in neighborhoods with SVI scores exceeding the national median, with highly concentrated poverty, lower levels of education, and a high proportion of Black residents (**Fig. 1C**). The inequities present in highly vulnerable neighborhoods likely reflect decades of disinvestment and structural racism, thus shaping risk in multiple ways, including through differential viral exposure (e.g., shared public transit, crowded housing, and less safe workplaces) as well as differential access to preventive tools (e.g., quality masks, vaccine access, indoor air quality) and health care interventions^13-18^. In addition, compared to residents of less vulnerable neighborhoods, people who lived in more vulnerable neighborhoods likely had poorer health, on average, prior to the pandemic – rendering them disproportionately susceptible to hospitalization or mortality once infected with SARS-CoV-2.

Second, it is likely that infections were undercounted in all neighborhoods. Mild infections may have been undetected, and as rapid antigen tests became more available and widely adopted, many at-home positive test results went undeclared to public health authorities^19^. Additionally, testing availability or testing site accessibility were often constrained, sometimes to a greater extent in the more vulnerable neighborhoods compared to the less vulnerable neighborhoods^20, 21^. What we observed based on confirmed cases reported to Philadelphia’s health department was a modest elevation in SARS-CoV-2 incidence among residents of more vulnerable neighborhoods, combined with more pronounced elevations in COVID-19 hospitalization and mortality (**Figs 2-3, Table 1**). If taken at face value, these results suggest that on average, SARS-CoV-2 infections tended to be more consequential among people living in the more vulnerable neighborhoods. This may be true, and another contributing factor could be that more cases were missed within the more vulnerable communities. Detection bias may have been particularly operative in vulnerable neighborhoods, many of which lack health care infrastructure, with residents facing barriers to care such as lack of transportation^20^ or paid time off. Test positivity, a gross indicator of testing adequacy, was persistently elevated in more vulnerable neighborhoods through most of the study period (**Fig 2B**)—lending credence to the hypothesis that COVID-19 incidence may have been differentially underestimated in more vulnerable neighborhoods. Thus, hospitalization and mortality rates may serve as more robust indicators of the cumulative impact of the COVID-19 pandemic.

Finally, deeply intertwined with the issue of social vulnerability are the contributions of racial inequity and racism to health, and this is particularly salient in Philadelphia. Compared to less vulnerable neighborhoods, Philadelphia’s more vulnerable neighborhoods -- which have high concentrations of Black residents (**Fig. 1F**) -- have unequivocally suffered greater total pandemic losses of life. Had residents of more vulnerable neighborhoods experienced COVID-19 mortality rates similar to those in more prosperous communities, almost 2,000 Philadelphians would still be alive. Our findings corroborate other recent investigations into the association between social vulnerability and COVID-19 incidence across the United States^22^, SVI and case fatality rates among counties nationwide^23^, and spatial inequities in COVID-19 impacts related to cumulative testing, positivity, confirmed cases, and mortality in Philadelphia at earlier points in the COVID-19 pandemic^24, 25^.

Even if we work now to close the racial gap in COVID-19 disease burden, we cannot reclaim the lives already lost, and the differential harms to Black Americans and vulnerable communities. In addition, nationally, between April 1, 2020 and June 30, 2021, COVID-19 mortality resulted in more than 140,000 children losing a parent or caregiver in the US, with children of racial and ethnic minority groups experiencing such a loss at 1.1 to 4.5 times the rate among non-Hispanic White children^26^. Caregiver deaths are likely to result in prolonged intergenerational social, economic, and health impacts. COVID-19 losses of life and health will reverberate for years to come.

### Strengths and Limitations

A key strength of our work is the use of the social vulnerability index, which is available for every census tract in the United States. Thus, this analysis may be replicated for communities across the country. These findings can help shape future public health policy and intervention strategies to be more equity-driven.

A key limitation of our work was the availability and quality of the data reported to the health department during an ongoing pandemic. As we have discussed above, cases were likely underreported as at-home tests became available. Reporting of COVID-19 hospitalizations also became complicated by individuals incidentally testing positive for COVID-19 during their hospital stay for a different cause (resulting in the terms hospitalized “with COVID” versus “for COVID”). These limitations in data have likely led to underestimations in the actual cumulative toll of the COVID-19 pandemic on communities.

Further, there may be residual confounding given that COVID-19 morbidity and mortality increase markedly with advancing age^27^, and it is thus reasonable to ask whether age differences across neighborhoods may account for the divergent health outcomes we observed across neighborhoods. There is no evidence that residents of the more vulnerable neighborhoods were older, on average, than the residents of less vulnerable neighborhoods. Philadelphia’s more vulnerable neighborhoods in fact skew younger, with 25.8% of residents under age 20, compared to only 12.6% of residents under age 20 in less vulnerable neighborhoods (**Fig 1E**). Additionally, a smaller proportion of residents were over age 65 in more vulnerable (13.5%) vs. less vulnerable (15.6%) neighborhoods **(Fig 1E**). Thus, this younger age distribution in the neighborhoods with high SVI scores should have provided protection against hospitalization or mortality from COVID-19. Given the well-documented disparate rates of COVID-19-related hospitalization and mortality by age ^28^ the results reported here are likely underestimates of the true impact of COVID-19 on Philadelphia’s more vulnerable communities.

## Conclusions

This analysis showed that COVID-19 has exacted an especially lethal toll in Philadelphia neighborhoods that scored high on the CDC’s Social Vulnerability Index. Therefore, in the context of COVID-19, the SVI was a useful tool for identifying neighborhoods at risk. In the ongoing COVID-19 pandemic, as well as during future public health crises, the SVI can inform a health-equity motivated approach, by allocating augmented resources and designing policies to protect the health and lives of vulnerable communities. Similar analyses should be replicated in municipalities across the US, to determine whether (and the degree to which) more vulnerable neighborhoods have suffered differentially from COVID-19, as well as to identify pandemic policies that mitigated or exacerbated inequities. Ultimately these investigations should inform a path forward, so that a person’s zip code does not undermine their health or life during epidemics, emergencies, or environmental threats.

## Data Availability

All data are available online. The 2018 Social Vulnerability Index Data was downloaded from the ATSDR/CDC site (https://www.atsdr.cdc.gov/placeandhealth/svi/data_documentation_download.html). We also used data from the 2020 American Community Survey (ACT) Five Year Estimates (available for download here: https://data.census.gov), and various COVID-19 data reported by the City of Philadelphia on the OpenDataPhilly website (https://www.opendataphilly.org/dataset/covid-cumulative-historical-data).

https://data.census.gov

https://www.opendataphilly.org/dataset/covid-cumulative-historical-data

## Acknowledgements

This work was funded by the Global Genomics and Health Equity Pilot Project RFA (GGHE-KP-2021-001).

## Notes

### Competing Interest Statement

The authors have declared no competing interest.

### Funding Statement

This study was funded by the Global Genomics and Health Equity Pilot Project RFA (GGHE-KP-2021-001).

### Author Declarations

All source data were openly available before the initiation of the study. The 2018 Social Vulnerability Index Data was downloaded from the ATSDR/CDC site (https://www.atsdr.cdc.gov/placeandhealth/svi/data_documentation_download.html). We also used data from the 2020 American Community Survey (ACT) Five Year Estimates (available for download here: https://data.census.gov), and various COVID-19 data reported by the City of Philadelphia on the OpenDataPhilly website (https://www.opendataphilly.org/dataset/covid-cumulative-historical-data).

